# ENCODE guided WGS analysis can identify trait associated regulatory regions driven by rare-variants

**DOI:** 10.1101/2024.11.06.24316407

**Authors:** Jack Thomas Flanagan, Kisung Nam, Seunggeun Lee

**Affiliations:** Graduate School of Data Science, Seoul National University, Seoul, South Korea; Genomic Medicine Institute, Seoul National University, Seoul, South Korea

## Abstract

Large-scale whole-genome sequencing (WGS) data provides unprecedented opportunity to explore the role of rare variants in non-coding regions on complex traits. However, unlike gene-based exome analysis, non-coding regions lack a well-defined unit for rare-variant testing. Here, we utilized 1,036,913 candidate Cis-Regulatory Elements (cCREs) from ENCODE as analysis units and conducted rare-variant association analysis for 100 traits with SAIGE-GENE+. In a discovery set of 150K White-British samples we identified 1,987 significant associations and replicated 88% of them in a validation set of 250K White-British samples. Associations were enriched in promoter-like signals (PLS) and proximal enhancer- like signals (pELS). Conditional analyses of 230 cCREs across five traits on GWAS variants identified 68 independent cCRE associations. A leukemia case study highlighted key loci, including a PLS for SRSF2 and a pELS for BCL6, demonstrating the effectiveness of cCRE- based analysis.

## Introduction

High-depth Whole Genome Sequencing (WGS) data present an unprecedented opportunity to explore the contribution of rare variants (MAF < 0.01) within non-coding regions to the heritability of complex traits. Recent large-scale sequencing efforts led by UK Biobank have cumulated in the public release of WGS data for 150,000, and subsequently 500,000 individuals, encompassing over 1 billion genetic variants ^1,2^. Previous UK Biobank data releases have included whole-exome sequences (WES) for 500,000 individuals, enabling in- depth analysis of coding variants^3^. However, approximately 90% of the associations discovered through genome-wide association studies (GWAS) are localized in non-coding regions^4^, highlighting the importance of non-coding genetic variants.

Despite the rapid expansion of WGS resources, an effective association analysis strategy for WGS data is not yet established. The single variant test, commonly employed in GWAS, has low power to identify rare variant associations. Unlike exome analysis, which is often conducted on a gene-based level, non-coding regions do not have a well-defined unit for set-based association tests^5,6^. Sliding-window approaches across the genome, with various functional information for within-set variant weighting, have been proposed ^7–9^. However, this broad, non-targeted approach, whilst comprehensive, may fail to capture discrete biological structures and potentially lose important biological context when defining variant sets.

In this study, we employ a targeted yet straightforward approach by using candidate Cis-Regulatory Elements (cCREs) from the ENCODE registry as the unit of analysis. At a fundamental level, cCREs implicated in association signals can be linked to potential roles as promoters or enhancers. This linkage is based on the expression patterns of key markers, including DNase, H3K4me3, H3K27ac, and CTCF, as well as their physical proximity to known transcription start sites. The ENCODE registry catalogues approximately one million cCREs, each spanning 150 to 350 base pairs, which exhibit distinct biological characteristics with the potential to influence gene regulation. By focusing on these meaningful biological structures within non-coding regions, our approach can enhance the understanding of how variation within these regions affect gene expression pathways, thereby influencing complex traits^8^.

We demonstrate that our approach can identify a large number of associations of rare regulatory variants. Utilizing the latest iteration of the rare-variant test, SAIGE-GENE+, we tested rare variants in cCREs for association across 100 traits and diseases in the UK Biobank. In a discovery set of 150,000 WGS White- British individuals, we identified 1,987 association signals and 1,334 associated unique cCREs^10^. We replicated 88% of them in a separate validation set comprising the remaining 250,000 White-British samples from the UK Biobank 500,000 WGS release. The associations were enriched in promoter-like signals (PLS) and proximal enhancer-like signals (pELS). Additionally, we performed comprehensive conditional analyses for 230 cCREs across five traits, resulting in 68 significant associations for rare variants. We also explored additional information from the ENCODE SCREEN Registry V3, highlighting potential enhancer/promoter-gene links and epigenetic marker expression profiles^11^. This placed association signals into a more informative biological context, notably in our analysis of loci identified in leukemia.

## Results

### Set-based association analysis targeting candidate Cis-Regulatory Elements (cCRE)

In contrast to whole-exome analysis, the analysis of WGS data presents substantial computational challenges and the absence of an established, interpretable unit of analysis within non-coding regions. To overcome this, our study leveraged the ENCODE project’s registry of candidate Cis-Regulatory Elements (cCREs) as a framework for grouping rare variants (MAF < 0.01) into variant sets^11^. The cCREs, which comprise 7.9% of the GRCh38 genome, provide a structured approach for categorizing non-coding variants. An overview of our analysis is provided in Figure 1.

**Figure 1:**
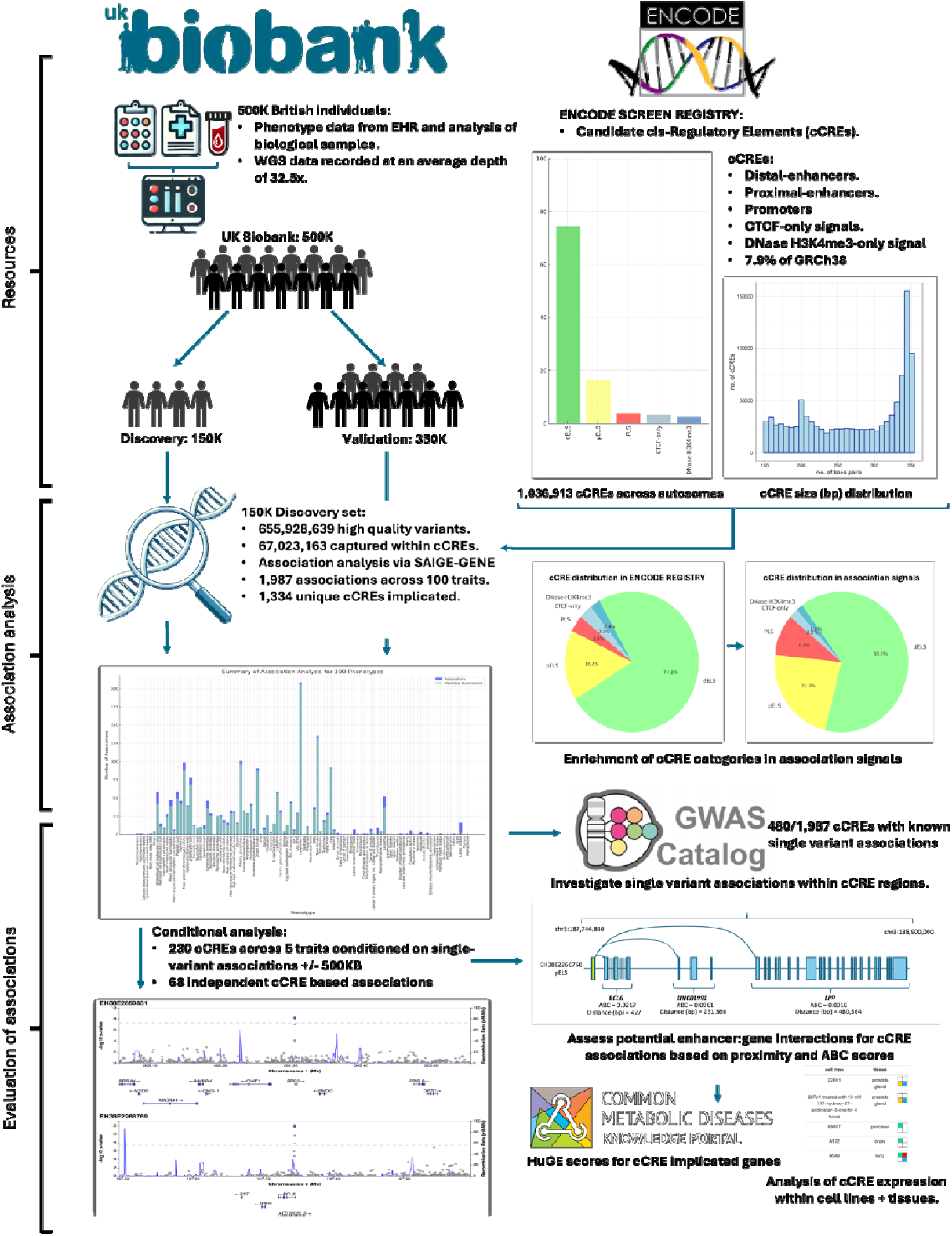
Overview of the analysis pipeline of our study including the UK Biobank and ENCODE resources.

We conducted a set-based association analysis across 100 phenotypes for White- British individuals within a cohort of 150,119 individuals, utilizing WGS data from UK Biobank. Each ‘set’ was defined by the start-end positions of a cCRE, totalling 1,036,913 sets encompassing 67,023,163 variants across chromosomes 1-22. Details on 100 phenotypes were shown in Supplementary Table S1. Association analysis for 100 phenotypes was performed using SAIGE-GENE+, applying a MAF threshold of 0.01.

Our analysis, summarized in Figure 2 and Supplementary Table S2, identified 1,987 genome-wide significant signals at *P* < 5.0 × 10^-8^ across 451 loci (based on genomic positions with a minimum of 500kb+/- between each locus), based on a Bonferroni correction for 1 million cCREs, implicating 1,334 unique cCREs. Among the 100 phenotypes analysed, association signals were detected in 72 phenotypes, whereas 28 phenotypes did not achieve genome-wide significance. Of these, 23 are binary traits and 5 are quantitative traits. Notably, genome-wide significant cCRE associations in binary traits were relatively rare, with only 116 out of the 1,987 signals observed, 52 of which were specific to leukemia.

**Figure 2:**
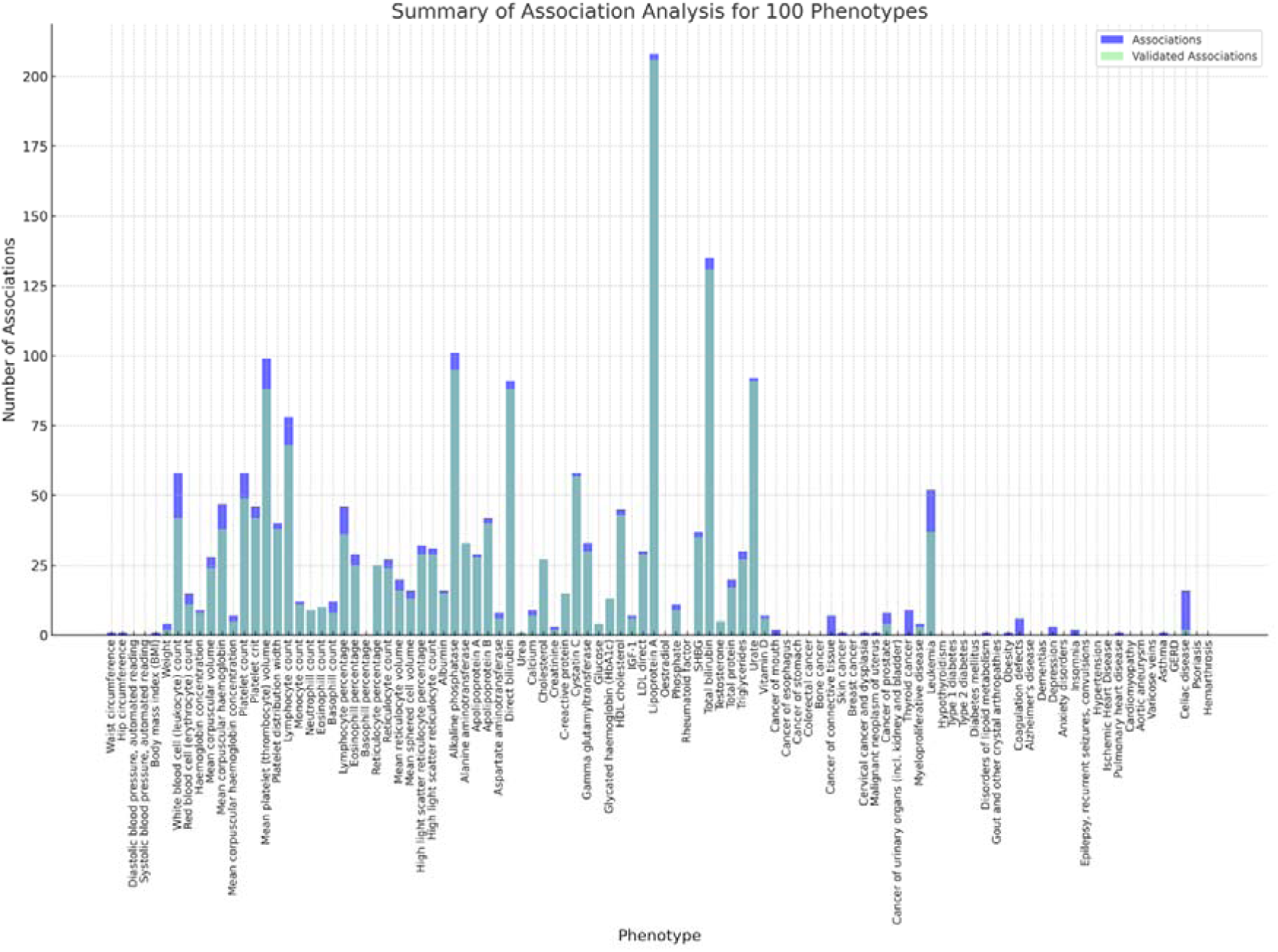
Summary of association analysis of one million cCRE-defined rare-variant sets across 100 traits recorded in UK Biobank. Associations identified in the 150K discovery set are plotted with the proportion of replicated signals from a validation cohort of 250K samples highlighted in green. Exact sample and case control numbers are recorded in supplementary table S1.

**Figure 3:**
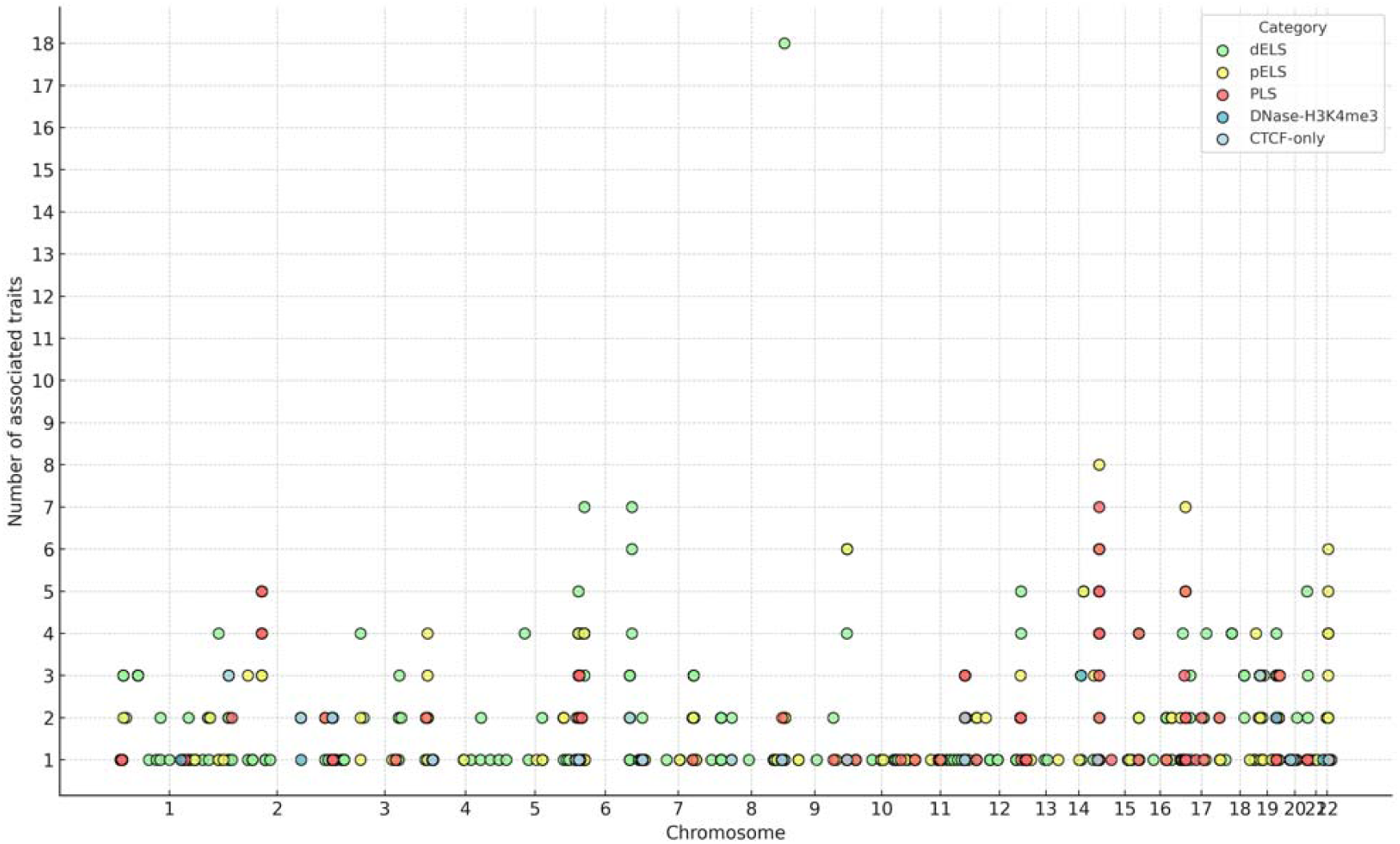
Classification and distribution of 1,334 cCREs reaching genome-wide significance across chromosomes 1:22 and the number of traits for which they exhibit association signals.

Quantitative traits in our analysis were grouped into three categories: physical measurements, blood count and blood biochemistry. In the category of physical measurements, we observed limited association signals, Waist circumference (1 signal), Hip circumference (1 signal), Diastolic blood pressure (0 signals), Systolic blood pressure (0 signals), Body mass index (1 signal). The only trait with more than one association signal was Weight with 4 signals distributed across 4 separate loci.

Traits in blood count and blood biochemistry exhibited a higher prevalence of cCREs reaching the genome-wide significance threshold (*P* < 5.0 × 10^-8^) with 40 of 54 traits across the two categories displaying 10 or more significant cCREs. However, some traits showed limited discovery, including Basophil percentage (0 signals), Urea (1 signal), Oestradiol (0 signals) and Rheumatoid factor (0 signals). In contrast, we observed many traits displaying extensive numbers of cCREs reaching genome-wide significance such as Alkaline phosphatase (101 association signals across five regions), Lipoprotein A (208 association signals distributed across a single region) and Total bilirubin (208 association signals across two regions).

Binary traits were categorized into 10 groups: Neoplasms, Endocrine/Metabolic, Hematopoietic, Mental disorders, Neurological, Circulatory system, Respiratory, Digestive, Dermatological and Musculoskeletal. Across these categories, the discovery of association signals was limited, with only 10 out of 40 traits exhibiting more than a single significant association. However, the association analysis for Leukemia was notably promising, with 52 cCREs spanning 7 regions being genome-wide significant.

Validation of association analysis results with 500K WGS release.

Validation of association signals identified in the 150K WGS cohort was conducted following the release of 500K WGS data by UK Biobank^2^ (Supplementary Table S2). This subsequent analysis focused exclusively on regions implicated in the initial 150K study, excluding samples from the interim 150K release to ensure independence. The analysis was again restricted to White-British samples. Of the 1,987 signals originally detected, 1,762 (88%) were validated at the genome-wide threshold of *P* = 5 x 10^-8^. Signals associated with quantitative traits showed high validation rates, with 91% (1,716/1,871) maintaining genome-wide significance in the validation cohort. In contrast, validation rates for binary traits were considerably lower, with only 39% (46/116) achieving genome-wide significance. Validation outcomes were consistent across quantitative traits, with no single phenotype showing disproportionate results. In binary traits, leukemia exhibited notably high validation rates (relative to other binary traits) with 37 of 52 associations confirmed, accounting for 80% of all validated binary trait signals. However, several traits exhibited particularly low validation rates, including thyroid cancer (0/9 validated), cancer of connective tissue (0/7 validated), coagulation defects (0/6 validated), and celiac disease (2/16 validated).

### Associated cCREs are enriched in PLS and pELS

In our analysis, cCREs were categorized into five distinct classifications: Promoter-like signatures (PLS), Proximal enhancer-like signatures (pELS), Distal enhancer-like signatures (dELS), DNase-H3K4me3 cCREs, and CTCF-only cCREs. These categories accounted for 3.8%, 16.2%, 74.2%, 2.4%, and 3.3% respectively of the 1,036,913 cCREs tested for association (Figure 4A). Among the cCREs achieving genome-wide significance, the distribution was as follows: PLS comprised 9.4%, pELS 22.7%, dELS 63.9%, DNase-H3K4me3 1.6%, and CTCF-only 2.2% (Figure 4B). These results indicate a significant enrichment of cCREs with PLS and pELS in our association analysis of 100 traits, with a statistically significant deviation (P = 3.81 × 10⁻³⁶) from the expected distribution of association signals across the five cCRE categories.

**Figure 4.**
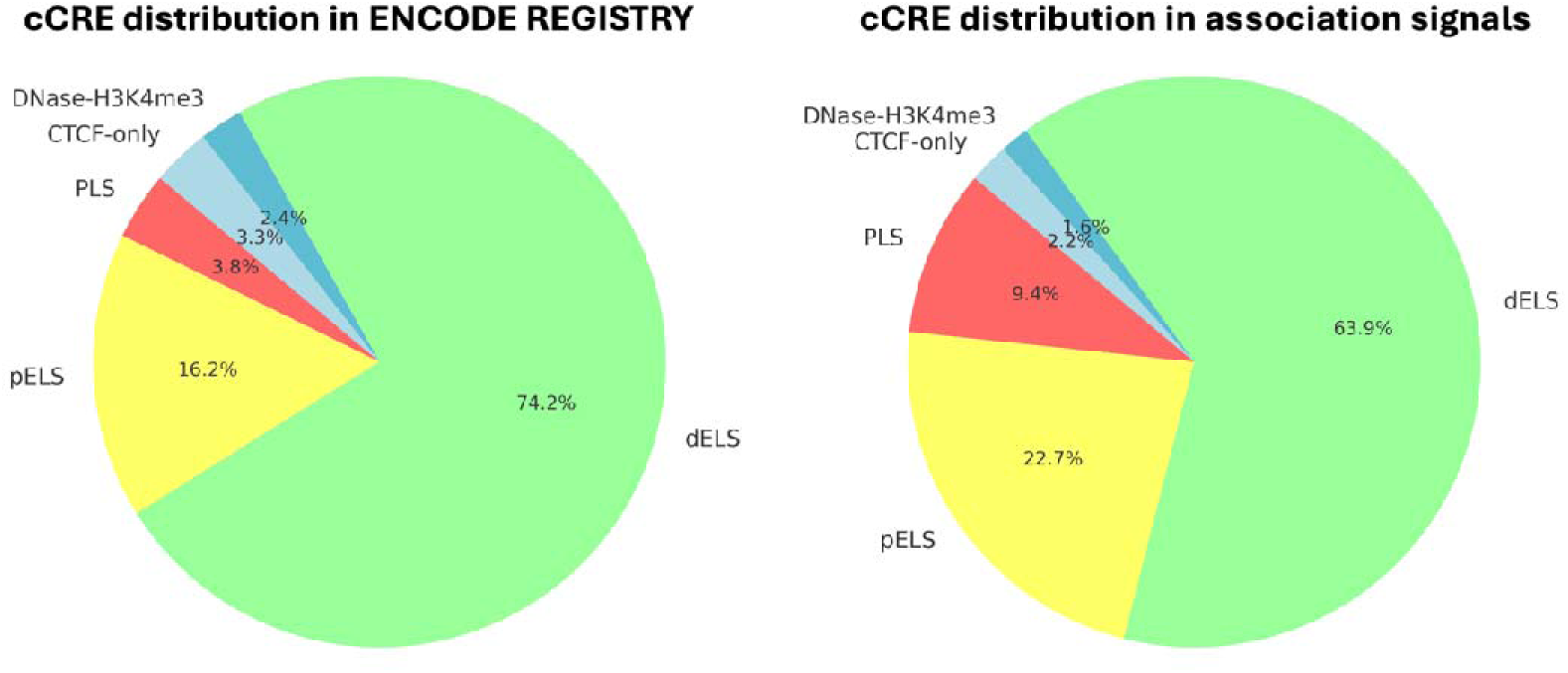
A: Proportion of each cCRE classification in the total ENCODE cCRE library. B: Proportion of each cCRE classification among the 1,334 reaching genome-wide significance for at least one trait of the 100 tested for association.

### Potential for cCREs exhibiting pleiotropic effects

Our results highlight several cCREs that show genome-wide significance across multiple traits, with 32 cCREs achieving significance across 5 or more different traits. cCRE EH38E1844080 (17: 7176933- 7177186), for example, is a pELS with strong association signals across 7 traits within the blood biochemistry category: Albumin, Alkaline phosphatase, Apolipoprotein B, Cholesterol, C-reactive protein, LDL direct and Triglycerides. cCRE EH38E1844080 is located within 2,505bp of the gene Asialoglycoprotein receptor 1 (*ASGR1*), a gene expressed exclusively in the liver with loss of mutations linked to decreased serum lipid levels^12^. Additionally, three other cCREs in close proximity, including a promoter region, displayed association signals across five lipid related traits, underscoring the potential to identify broader sections of non-coding regions that exert influence over multiple traits.

### Conditional analysis on GWAS identified loci

To explore the potential for the novel association discovery, we conducted conditional analyses for five traits: Leukemia, HDL cholesterol, white blood cell (leukocyte) count, urate, and cystatin C. Using GWAS summary statistics from the Neale Lab for the UK Biobank imputed dataset (N=361,194 imputed up to the Haplotype reference consortium, UK10K and 1000 Genomes Project), we identified genome-wide significance SNPs located within 500kb of a cCRE-defined association signal from our results. In regions where single variant associations were present within 500Kb of a cCRE, up to 10 variants were included in a series of stepwise conditional analyses for the corresponding locus. The selection of variants was based on the five strongest signals in both positive and negative directions. The results are presented in Supplementary Table S3.

In the conditional analysis for leukemia, 14 of the 23 cCREs reaching the threshold for genome-wide significance were located within 500Kb of a single variant identified in the Neale lab GWAS summary statistics (reference for Neale lab). After conditioning on 7 variants, all 14 cCREs across 4 loci retained genome-wide significance. For HDL cholesterol, 44 out of 45 cCREs that reached genome-wide significance across 10 loci were also within 500Kb of a single variant association signal. After conditioning these 44 cCREs on 100 variants, 16 retained genome-wide significance across 9 loci. In the analysis of white blood cell (leukocyte) count, 26 cCREs were conditioned on 43 SNPs, with 3 retaining genome-wide significance, and genome-wide significant signals were confined to 2 out of 5 originally identified loci. For urate, conditional analysis on 92 cCREs across 6 loci using 32 variants from the Neale lab summary statistics resulted in 23 cCREs retaining genome-wide significance across 4 loci. Lastly, in the analysis of cystatin C, 56 cCREs identified across 5 loci were conditioned on 47 variants, with 14 cCREs retaining genome-wide significance across all loci post conditional analysis.

Figure 5 highlights a key locus on chromosome 20 with a particularly strong association with Cystatin C. In this region we observe a cluster of cCREs displaying strong association signals within close-proximity to the gene *CST3,* the gene encoding the protein cystatin C. Following conditional analysis, nine cCREs retained genome-wide significance including four dELS, three pELS, one DNase-H3K4me3 and one PLS. These findings suggest that variations within these regulatory regions may significantly influence serum Cystatin C levels.

**Figure 5:**
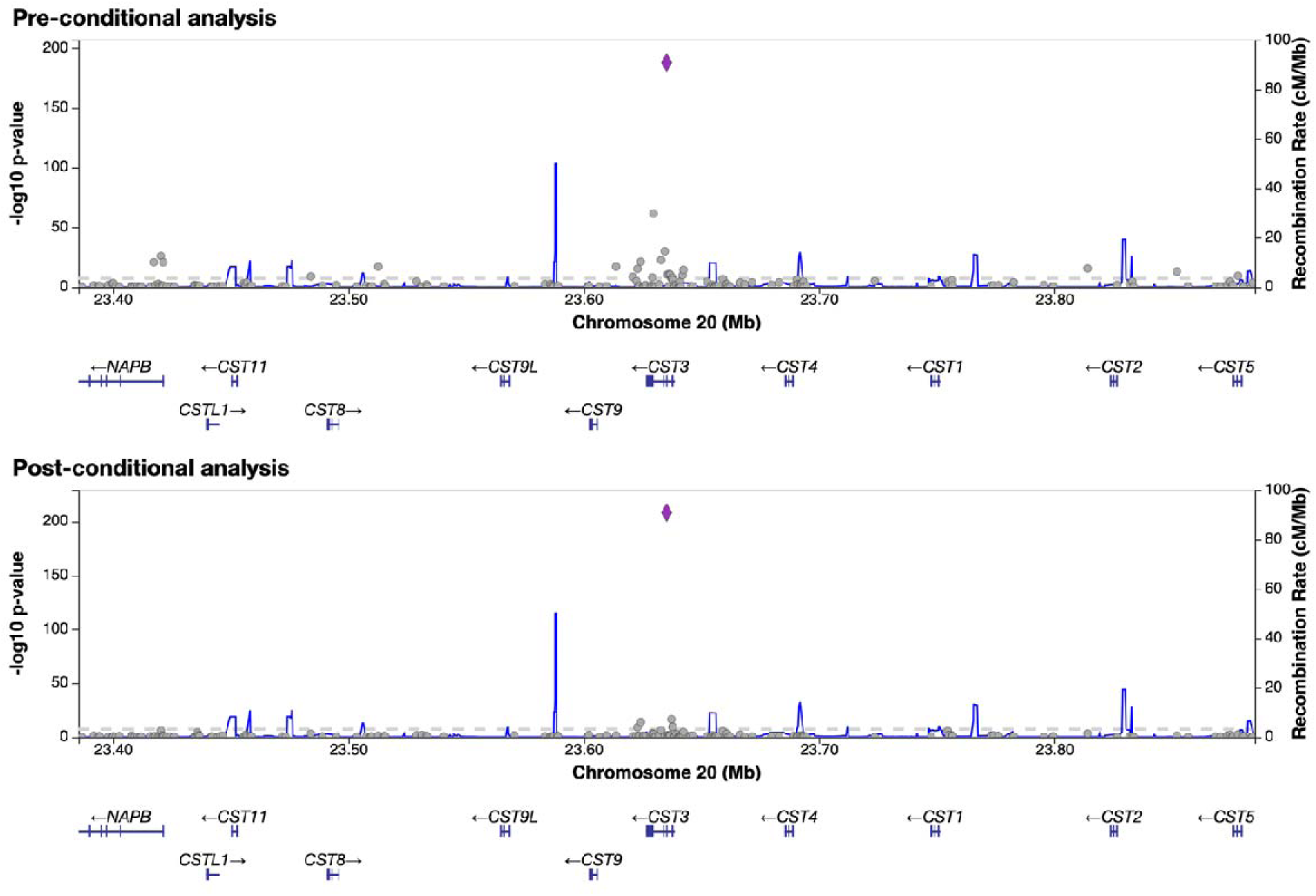
Locus-zoom plots for the Cystatin C associated locus with the lead association signal driven by the dELS cCRE: EH38E3424825 (P = 1.51 x 10) and highlighted by the purple diamond. Here we present associated signals in the region (plotted on the middlepoint of the cCRE signal) pre and post-conditioning on single variant association variants in the region (+/-500kb).

### Investigating potential enhancer/promoter-gene links

In our downstream analysis of cCREs post-conditional analysis, we observed multiple instances where evidence from the SCREEN Registry V3 supports their role in the expression of a given trait (Supplementary Table S4). For Leukemia we observe the cCRE EH38E3244983, a PLS with a strong association signal in both discovery (*P* = 7.7 x 10^-20^) and validation (*P* = 1.6 x 10^-47^) sets located in close proximity (< 500bp) to the gene *SRSF2* (Serine And Arginine Rich Splicing Factor 2). *SRSF2* was implicated in our whole-exome sequencing analysis (*P* = 3.9 x 10^-65^, N cases = 3,075) and has been linked to acute myeloid leukemia, with increased frequencies of mutations observed in individuals with the disease. These mutations are linked to aberrant splicing and disruption of additional genes related to acute myeloid leukemia^13,14^.

Furthermore, we observed a grouping of nine pELS cCREs in the 22q11.22 region, the immunoglobulin lambda locus. These cCREs spanned the region of 22:22686850-22900803, with the lead signal defined by cCRE EH38E3470428 (*P* = 9.8 x 10^-53^, validation *P* = 2.0 x 10^-98^) and in proximity to a collection of genes including *IGLL5*, *MIR5571*, *IGLJ1*, *IGLC1* and *IGLV3-1,* all within 20,000bp. *IGLL5* showed a strong association signal in our WES analysis for leukemia (*P* = 4.3 x 10^-64^) with frequent mutations observed in chronic lymphocytic leukemia cases^15^. On chromosome 3, we identified additional three pELS cCREs, two of which were replicated in our validation set. These potential enhancers were located within 1000bp of the gene *BCL6*, a transcription repressor linked to high expression levels in acute myeloid leukemia cell lines and associated with cell proliferation and survival. Inhibition of *BCL6* expression has been linked to reduced cells proliferation and increased chemotherapy sensitivity^16^. Additionally, we identified pELS cCRE EH38E2859331 on chromosome 1, located within 432bp of the gene *BTG2*, an anti-proliferation gene with potential tumour-suppressing qualities, frequently mutated in B-cell malignancies^17^.

In addition to investigating the relationships between cCREs and their proximal genes, we considered methods to assess the potential for enhancer regions to interact with multiple genes over larger genomic distances. We utilised the CATLAS resource developed by Zhang et al 2021, providing comprehensive data for potential enhancer-gene interactions across 111 cell types using the activity-by-contact (ABC) model^18,19^. Analysis was completed on a subset of cCREs that both retained genome-wide significance post-conditional analysis and overlapped with the CATLAS library of cCREs. We observed 16 cCREs across 4 traits that exhibited ABC scores meeting a 0.015 threshold, suggesting potential gene targets (Supplementary Table S6). These ABC scores provide preliminary evidence that some cCREs interact with multiple genes and these interactions are not limited to the most proximal gene. These results also highlight stronger ABC scores with more distant genes with the potential for interactions extending over 1MB. In our analysis of leukemia, we observed two pELS cCREs EH38E2266768 (*P* = 1.92E-13) and EH38E2266769 (*P* = 1.61E-16), which, according to the ABC model, influence multiple distal genes in addition to *BCL6*, the most proximal gene. In fact, this model predicts that these cCREs may exert a greater influence over the expression of *LINC01991* (ABC = 0.09614447) and *LPP* (ABC = 0.0915) in comparison to *BCL6* (ABC = 0.0217278).

To supplement the biologically relevant information for the cCREs implicated in association analysis, we assessed signals of DNase, H3K4me3, H3K27ac, and CTCF for cCREs in over 1,500 cell types in the ENCODE SCREEN registry v3 (Supplementary Table S4). For example, PLS EH38E3244983 on chromosome 17 showed the strongest signals across DNase, H3K4me3, H3K27ac, and CTCF in the GM12878 cell line, a lymphoblastoid cell line derived from lymphocytes, with a DNase z-score of 3.62 (1.64 representing the 95th percentile and the recommended threshold for filtering based on activity). The second and third strongest signal profiles were observed in OCI-LY7 and K562 cell lines, representing B-cell lymphoma and chronic myeloid leukemia models, respectively. These insights provide further evidence for identifying true association signals and linking them to biological pathways and mechanisms impacting trait heritability.

## Discussion

This study demonstrates that ENCODE cCREs offer an effective framework for analysing WGS data. By leveraging cCREs to group rare variants, we identified numerous association signals across 100 traits. Additionally, our approach addresses the challenge of interpreting association signals in non-coding regions by linking genetic variants to well-defined biological structures, thereby elucidating the mechanisms through which these variations impact trait heritability. With supporting data indicating potential roles as promoters, proximal or distal enhancer regions, in tandem with GENCODE annotation data concerning transcription start sites, we can connect association signals in non-coding regions to candidate target genes^20^. Summary statistics for our cCRE-based association analysis of 100 traits in UK Biobank have been uploaded to a Pheweb-like server to visualise associations using an adapted version of the Pheweb software .

The results from our WGS association analysis of 100 traits highlighted a substantial disparity in performance between quantitative and binary traits. Case/control ratios were heavily imbalanced in our discovery set and many traits reported less than 1000 cases. Whilst resources such as the UK Biobank provide unique access to the largest collections of WGS data, sampling methodology is designed to reflect the general population and not to specifically target individuals with one particular trait^22^. Consequently, capturing samples for specific diseases is limited, especially for uncommon and rare diseases. Furthermore, these limited case numbers for binary traits in both discovery and validation sample sets negatively impacted the replication of association signals. Despite limitations, we identified several cCREs that achieved genome-wide significance in binary traits and were successfully replicated in our validation sets. Analyses for prostate cancer, leukemia, and myeloproliferative disease revealed multiple instances of replicable genome-wide associations. Most of these associations showed stronger signals (in terms of *P* value) in the larger validation sets.

Our analysis revealed a number of regions containing a high density of cCREs exhibiting strong association signals. On chromosome 14 we observed a series of 14 cCREs showing association with 5 or more traits. This particular region of interest was observed in traits related to the immune system, such as white blood cell count and leukemia, located between 14:105745623-106851551. This region comprises over 100 genes responsible for the heavy chain of antibodies and is recognized as one of the most complex and variable regions of the genome^23^. Furthermore, this extreme complexity is linked to large numbers of structural variants in the region, which can lead to difficulties in reliably genotyping variants^24,25^. Within this region, we assessed sequencing depth for variants implicated by cCREs and observed generally poor genotyping quality, leading us to exclude cCREs in this region from downstream analysis.

Comprehensive conditional analysis of cCREs in five example traits highlighted the utility of capturing rare and ultra-rare variant sets, as we identified multiple regions of association signals that had not been captured by single-variant analysis of the UK Biobank imputed sample sets. These signals remained independent and genome-wide significant following multiple rounds of conditional analysis demonstrating the capability of cCRE based rare-variant analysis to identify novel associations. Furthermore, these novel associations can be supplemented with enhancer-gene interaction studies such as those recorded in CATLAS. The identification of potential target genes gives an additional layer of depth to the results of cCRE based association studies and can provide a greater insight into the mechanism by which these signals may be implicated in the heritability of a given trait.

Whilst we observed novel sources of association stemming from cCREs, it is important to consider potential limitations in statistical power due to low MAF ranges targeted in our analysis. Additionally, sizes of cCREs are relatively small (only 150 to 350 base pairs), whereas gene-based regional analyses typically cover regions measured in kilobases. One strategy to increase the power is to aggregate multiple cCREs into larger units of analysis. Methods for gene-set analysis, such as the Gene-set analysis Association using Sparse Signals (GAUSS), can combine association signals of multiple genes, offering improved capabilities to identify associations ^26^. Adapting these methods for cCRE-based variant-set analysis could help mitigate the power limitations associated with focusing on small genomic regions.

Here we have presented a comprehensive evaluation of using cCREs to specifically interrogate non-coding regions of WGS data through rare variant set-based association analysis. Our analysis encompassed a wide range of complex traits and diseases, demonstrating a consistent ability to identify association signals from rare and ultra-rare variants, taking full advantage of the information provided for variants with extremely low MAFs in WGS data. We then highlighted the validity of these signals through replication, validating 88% of the association signals in a separate cohort. Using data from ENCODE, we highlighted the capacity to link cCREs to complex traits through cCRE classifications, potentially linked genes, and expression data for DNase, H3K4me3, H3K27ac, and CTCF, indicating activity in specific cell lines. The ENCODE library of cCREs offers invaluable data for effectively targeting non-coding regions and gaining insights into the functional impact of mutations outside gene-coding regions of the genome.

## Methods

### Data processing and study cohort

The sample set used in this study was sourced from the UK Biobank release of WGS data for 500,000 individuals. Sequencing was performed for 490,640 UK biobank participants using the Illumina NovaSeq 6000 platform. Average coverage across the entire sample set is 32.5x, with a per individual average coverage of 23.5x. Full details of sampling and sequencing methodology are reported by UK Biobank^1,2^. Analysis was performed on the GraphTyper^27^ population level WGS variants (N=655,928,639), stored in variant call format (VCF) and divided into 50KB intervals. Genotype files were prepared with bcftools (v1.15.1)^28^ and multiallelic sites were split in preparation for analysis using SAIGE-GENE+. Genotype files were further processed using plink2 (v2.00a3.1LM)^29^ for concatenation of chunked files and conversion to the PLINK binary file format for analysis. The total WGS sample set was limited to ‘White-British’ individuals and variants filtered to include only those found within regions defined by candidate cis-regulatory elements (cCREs). Accession codes, start/end positions (hg38) and annotation information for 1,063,878 Candidate cis-regulatory elements were sourced from the ENCODE Registry of cCREs (Registry V3)^11^ via screen.encodeproject.org. Filtering of 655,928,639 variants captured 67,023,163 across 1,036,913 cCREs spanning the autosomes.

### Association analysis for 100 traits in UK Biobank

Phenotype data for UK Biobank is sourced from extensive per sample baseline data, electronic health records, blood, urine, and saliva analysis, including biochemical markers among other measurements, as fully reported by UK Biobank^30^. We selected 40 binary traits (based on the International Classification of Diseases, Tenth Revision (ICD10) codes) and 60 continuous traits based on measurements in UK Biobank (Full details regarding traits, per trait sample size, and case/control numbers reported in Supplementary Table S1).

Association analysis for 100 traits was performed using SAIGE-GENE+ (v2.0.2). SAIGE-GENE+ supports efficient variant-set based analysis of large-scale data sets, collapsing ultra-rare variants (MAC < 10) within each set into single units, performing multiple tests (Burden, SKAT and SKAT-O) to accommodate varying effect directions within variant-sets, whilst supporting multiple MAF thresholds and providing a final ‘Cauchy’ combined *P* value to summarise. Group files to establish units of analysis were defined by start:end positions of cCREs. Prior to association analysis, a sparse genetic relationship matrix was created and used for SAIGE-step1 to adjust for sample relatedness. We also incorporated age, sex, batch and the top 10 principal components as covariates. We applied MAF thresholds of 0.01, 0.001, and 0.0001, inverse normalisation for quantitative phenotypes, and saddlepoint approximation for binary phenotypes to account for imbalances in case/control ratios.

Association analysis was performed on a discovery set of 150,000 WGS samples from the UK Biobank interim WGS release, with the genome-wide significance threshold of *P* = 5 x 10^-8^ based on a Bonferroni correction accounting for ∼1 million cCREs tested for association. To validate the association signals, we repeated our analysis using a replication sample set using the remaining 250,000 ‘White-British’ samples of the 500K WGS release. We targeted loci where associations were identified in the discovery set and used a threshold of *P* = 5 x 10^-8^. To determine cCREs exhibiting genome-wide significance, we used the Cauchy combined test statistic *P* value (results for cCREs exhibiting genome-wide significance alongside sample size and case/control ratio for each trait are found in Supplementary Table S2).

### Conditional analysis

We selected five traits (HDL cholesterol, White blood cell count, Urate, Cystatin C and Leukemia) for the conditional analysis. To identify variants on which to condition, we used the Neale lab GWAS results for 361,194 UK Biobank samples imputed up to the 1000 Genomes and Haplotype reference consortium reference panels. Summary statistics for these traits were converted from GRCh37 to GRCh38 and single variant associations within ±500KB of a cCRE-led association signal were selected for conditioning. When multiple single-variant associations present in the same locus, we selected the top 10 strongest associations (based on T-statistic) for both positive and negative effect directions. Due to limitations in the number of variants that can be included in the model for conditional analysis with SAIGE-GENE+ during step 2, we directly incorporated genotypes for these variants into step 1 of the SAIGE-GENE+ process. Genotypes were added into the phenotype file as covariates in step1 of the SAIGE GENE+ process. Locuszoom plots were generated for each locus subject to conditional analysis, highlighting genes in close proximity to cCRE association signals.

### Identification of single-variant signals within cCRE regions

To search for evidence of single-variant associations originated from cCRE, we queried the GWAS catalogue using the gwasrapidd R package. Start:end positions for each cCRE with *P* < 5 x 10^-8^ and the associated trait were used to search the GWAS catalogue for evidence of known single-variant associations that directly implicate cCREs in our own results. Instances of single-variant associations within cCRE defined regions are reported in supplementary table S5.

### Gene – enhancer/promoter links

cCREs that retained genome-wide significance (*P* = 5 x 10^-8^) post-conditional analysis across HDL cholesterol, White blood cell count, Urate, Cystatin C and Leukemia traits were subject to further analysis using SCREEN REGISTRY V3. For each cCRE, we report the closest gene (in base pairs) and the cell line in which overall expression signals of the four following epigenetic markers is the strongest: DNase, H3K4me3, H3K27ac and CTCF (supplementary table S4). Expression signals are based on Z-scores recorded in SCREEN REGISTRY V3, with a Z-score of 1.64 and above representing the 95^th^ percentile. Information concerning activity across cell lines and nearby genomic features was used to guide a literature search to investigate the relationship between a given trait and the potential gene-target of the associated cCRE. In addition to assessing the proximal genes for post-conditional analysis cCRE set we investigated the potential interactions with distal genes based of evidence in the CATLAS database of enhancer-gene interactions based on the ABC model. We compared the two sets of cCREs (ENCODE and CATLAS), looking for instances of overlaps between the two sets to finalise a list of ENCODE cCREs that had both been subject to conditional analysis and also overlapped with those found in CATLAS. Using a minimum threshold of ABC = 0.015, we documented potential enhancer-gene interactions for our target set of cCREs. To further assess the relevance of the cCRE-implicated gene to the target trait, we used the Human Genetic Evidence Calculator (HuGE) scores, calculated from the combined bayes factor of common and rare variation to determine the level of evidence for a given gene-trait relationship.

### Whole-exome sequencing analysis

In addition to WGS, we performed gene-based association analysis using SAIGE-GENE+ for 100 traits on whole-exome sequencing data, in a cohort of 393,247 White British individuals in UK Biobank. We excluded monomorphic variants and applied filters for missingness rates of > 0.1 and HWE P value < 10^-15^. We generated group files for SAIGE-GENE+ to define the variants in each gene and their accompanying functional annotation data. Group files were generated using the loss-of-function transcript effect estimator (LOFTEE). We classified a variant as loss-of-function (LoF) only if it was labeled as a high-confidence (HC) LoF variant, while variants with low-confidence (LC) were regarded as missense variants.

## Data availability

The analysis results for 60 quantitative and 40 binary phenotypes of UKB WGS data analysis results are available at a pheweb-like server which can be accessed at https://ccre-ukb-150kwgs.leelabsg.org/

## Code availability

PheWeb, an open-source tool to build a website to browse GWAS, was adapted to cater for cCRE based association analysis, code is available at https://github.com/leelabsg/cCRE-site-ukbb. SAIGE GENE+ is an open-source R package available at https://github.com/saigegit/SAIGE.

## Author Contribution

J.F. and S.L. designed experiments. J.F. performed experiments and analyzed the UKB WGS data. K.N adapted PheWeb software to develop a PheWeb-like server for cCRE association analysis results for 100 traits. K.N performed analysis of UKB WES data. J.F. and S.L. wrote the manuscript.

## Acknowledgements

This research was supported by the Brain Pool Plus (BP+) Program through the National Research Foundation of Korea (NRF) funded by the Ministry of Science and ICT (2020H1D3A2A03100666). This research was conducted using the UKBB Resource under application number 45227.

